# A Delphi Process to Determine the Bellwether Procedures for Trauma Systems Globally: A Study Protocol

**DOI:** 10.1101/2025.02.11.25322065

**Authors:** MF Bath, T Edmiston, J Amoako, A Ratnayake, D Bagaria, R Menon, J Wohlgemut, M McKenna, K Bateman, K Hancorn, J Shepherd, S Yoong, L Hobbs, BG Smith, C Whiffin, TC Hardcastle, A Conway Morris, T Weiser, T Bashford

## Abstract

**Introduction:** Traumatic injuries are responsible for a huge amount of mortality, morbidity, and disability globally. Within global surgery, Bellwether procedures have previously been identified to measure the surgical proficiency of a hospital or a region, however traumatic injuries often have distinct epidemiological and demographic patterns, compared to routine surgical care. Using a focused set of procedures or processes to measure trauma performance could allow for improved trauma assessments and outcomes on a global scale.

**Methodology:** An international Delphi study will be conducted in attempt gain consensus on the optimal procedures and processes that can be used to assess the performance of trauma care within any region or hospital worldwide. Recognised guidelines for conducting the Delphi process will be followed, comprising of 3 separate rounds and participation from a public and patient involvement (PPI) group. Respondents will be identified through pre-existing collaborative networks and research partners, to ensure adequate global representation from across the trauma care pathway. The final Bellwether procedures will be those that have shown consensus in agreement and in stability throughout successive rounds

**Discussion:** This Delphi process aims to identify the optimal Bellwether metrics that could be used to assess trauma care worldwide. Using a focused set of procedures or processes to assess trauma performance globally will reduce complexity and improve ease of use compared to current methods.

## Introduction

Injuries and violence are responsible for an estimated 10% of all years lived with disability worldwide (1). Trauma contributes to the greatest loss of disability-adjusted life-years for young adults worldwide (2), resulting in a huge strain on healthcare services and regional economic outputs (3,4). In response to this, many countries have implemented trauma systems to improve their trauma response and many have demonstrated overall improvements in mortality rates following implementation (5–8). The majority of documented trauma systems have been implemented within High-Income Countries (HICs), yet their designs are often non-transferable to the Low- or Middle-Income Country (LMIC) setting, where the need is greater. As trauma systems are often embedded within existing emergency services, such systems are weaker where the pre-existing health systems are fragile or suffering from conflict or humanitarian disaster.

Despite the efforts of agencies, such as the World Health Organisation (WHO) through their International Registry for Trauma and Emergency Care and regional trauma initiatives (9), it remains challenging to understand existing trauma system performance, compare systems with other settings, and plan targeted interventions to improve outcomes. Using a focused set of procedures or processes to assess trauma performance could lead to improved measures, both from ease of use and reduced complexity. In global surgery, the Bellwether procedures of Caesarean delivery, laparotomy, and treatment of open fracture are closely associated with the ability to perform all obstetric, general, emergency, and orthopaedic procedures (10). They represent core procedures that all global citizens should expect access to and are indicative of an effective surgical system; as an agreed global denominator (11), Bellwether procedures support the assessment and monitoring of access to timely surgery in hospitals and healthcare systems across cultural and political boundaries, and are currently utilised by multiple countries worldwide across a range of resource settings (12–16).

However, trauma-related injuries have a distinct epidemiological pattern, compared to general surgery cases, with the added complexity of time dependency. To date, no Bellwether procedures or processes have been defined for trauma, limiting the ability to benchmark different trauma systems across different contexts. The use of a Delphi process has previously been implemented to good effect within trauma and global surgery, such as by the BEACON Collaborative in defining textbook outcomes for trauma and non-trauma emergency laparotomy (17), or by the NIHR Global Research Health Unit on Global Surgery in developing global guidelines for emergency general surgery (18). Building on this experience, our aim is to perform an international Delphi study to produce consensus Bellwether procedures that can be used to assess the performance of trauma systems worldwide.

## Methodology

The study design will follow published and cited guidelines for Delphi process methodology (19), divided into 1) Identifying the Problem Area 2) Panel Member Selection 3) Delphi Rounds 4) Closing Criteria.

### Identifying the Problem Area

Current initiatives to benchmark quality of care across global trauma systems remain inadequate. A potential solution is to identify Bellwether measures within trauma care that are indicative of overall system performance (20). These include both specific surgical procedures to manage traumatic injury, such as a trauma laparotomy, and system processes within the trauma care pathway, such as diagnostic imaging.

We will define Major Trauma as a “significant injury or injuries that have potential to be life-threatening or life-changing sustained from either high energy mechanisms or low energy mechanisms in those rendered vulnerable by extremes of age” (21). This broad definition of major trauma allows the focus to shift away from a purely mortality-focused perspective trauma outcomes, to a more holistic definition which takes account of morbidity and rehabilitation potential (22).

An initial list of potential measures will be compiled through a combination of systematic literature searching, expert opinion, and discussion with key stakeholders in trauma care from across the globe, identified through contacts of the International Health Systems Group, the Moynihan Academy, and supporting partners. This list will be used to inform the Delphi rounds.

### Panel Member Selection

Involvement of the panel members involved in the Delphi process will be performed through a purposive snowballing technique, using pre-existing collaborative networks and research partners. Given the broad perspectives required to answer the research question, a high heterogeneity in the breadth of trauma care experience among the panel members is required. We will aim to cover any healthcare worker globally involved in the care of trauma patients, from Pre-Hospital and Emergency Medicine through to Surgery and Intensive Care.

Based on similar work (17), we would hope to recruit between 300-400 initial respondents, with the expectation of moderate attrition between rounds. Despite this heterogeneity, we expect to generate consensus within the three rounds of the process given the pre-existing published work in characterising traumatic injury.

### Delphi Rounds

Three separate sequential rounds in the Delphi process are planned for, to be conducted using a secure online platform. Participants will be asked to rank the varying measures suggestive using a 5-point Likert scale, with only collectively agreed measures proceeding to the next round. The number of options available will be narrowed down in sequential rounds. An option for anonymous comments to be submitted at the first stage will be included, with any comments showing sufficient consensus across respondents will be added at subsequent rounds.

Participants will be masked to each other’s identity and responses, and all analysis undertaken also masked to the participants. However, contact details will be recorded centrally to allow for appropriate distribution of subsequent rounds of the exercise to relevant individuals. Prior to the final round of the Delphi, a public and patient involvement (PPI) group will be convened, comprised of lay individuals with specific experience within major trauma. This will be undertaken to better inform the final round of the Delphi and will be reported against GRIPP2 short form as appropriate (23).

### Closing Criteria

Given the relative heterogeneity of the participants, based on similar work, consensus for a given statement in initial rounds will require a median score of >3.5 with an interquartile range (IQR) of ≤1, and show stability across subsequent rounds (19). The final Bellwether procedures will be those that have shown consensus in agreement and in stability throughout successive rounds (19).

### Ethical Approval

Ethical approval has been obtained from the Engineering Research Ethics Committees at the University of Cambridge (application no 525). The study will be led through a collaboration between the Moynihan Academy (www.moynihanacademy.org.uk) and the International Health Systems Group (https://www-edc.eng.cam.ac.uk/research/international-health-systems), based at the University of Cambridge.

## Discussion

The assessment of effective surgical systems through surgical Bellwether procedures has revolutionised global surgical research and policy (11). However, the burden of major trauma to populations and economies globally is vast and ensuring an equivalent approach is key if optimal trauma care strategies and systems are to be identified and evaluated. This Delphi process aims to identify the necessary trauma Bellwether measures within trauma care, by utilising an international cohort of stakeholders from specialities across the trauma pathway.

The care of major trauma patients requires multiple aspects of healthcare to integrate effectively and does not lie solely within surgical care. Optimal trauma care requires co-ordination between pre-hospital, anaesthesia and intensive care, surgical teams, and rehabilitation (24), with provision from supporting services, such as radiology and blood transfusion. In respect of this, ensuring the options available for the initial rounds of the Delphi process are both procedures (e.g. a trauma laparotomy) and processes involved within trauma care (e.g. provision of blood transfusion) is essential.

Completion of this Delphi process will allow for important measures of trauma care to be identified that will have applicability in trauma system assessment, both in terms of mortality and morbidity outcome and in economic measures.

## Data Availability

Study protocol only

## Declarations

### Ethics approval

Ethical approval was received from the University of Cambridge Department of Engineering’s Research Ethics Committee (application no 525)

## Acknowledgements

We are grateful for the support from the Association of Surgeons of Great Britain and Ireland (ASGBI), Asian Collaboration for Trauma (ACT), Global Anaesthesia, Surgery and Obstetric Collaboration (GASOC), Primary Trauma Care (PTC) Foundation, and NIHR Global Health Research Group on Acquired Brain and Spine Injury (GHRG-ABSI)

